# Breakthrough infection with SARS-CoV-2 and its predictors among healthcare workers in a medical college and hospital complex in Delhi, India

**DOI:** 10.1101/2021.06.07.21258447

**Authors:** Pragya Sharma, Suruchi Mishra, Saurav Basu, Neha Tanwar, Rajesh Kumar

## Abstract

**Introduction:** The study objective was to determine the breakthrough infection rate of Covid-19 (SARS-CoV-2) infection in those vaccinated with either BBV152 or AZD1222 (ChAdOx1-S) vaccine among healthcare workers (HCWs).

**Methods:** A cross-sectional analysis was conducted a medical college and hospital complex in Delhi, India through telephonic interviews among HCWs who had received at-least one dose of a Covid-19 vaccine during January to March’ 2021. Breakthrough infections were operationally defined as occurrence of Covid-19 infection ≥14 days after administration of two doses of either Covid-19 vaccine.

**Results:** We enrolled 325 HCWs with mean (SD) age of 29.1 (9.9) years including 211 (64.9%) males. Two seventy nine (85.8%) HCWs were fully vaccinated while 46 (14.2%) were partially vaccinated. There were 168 (51.7%) BBV152 and 157 (48.3%) AZD1222 (ChAdOx1-S) recipients.

A total of 37 (11.3%, 95% C.I. 8.3, 15.3) breakthrough infections were observed in the HCWs. The median (IQR) time until incidence of Covid-19 breakthrough infection since receiving second dose of either Covid-19 vaccine was 47 (28.5, 55) days. Additionally, 20 (6.1%) non-breakthrough Covid-19 infections were recorded in the HCWs post vaccination with either a single dose of a Covid-19 vaccine or both doses but prior to a period of 14 days since administration of the second dose.

Most breakthrough infection cases (94.4%) were mild and did not require supplemental oxygen therapy. HCWs without a history of natural Covid-19 infection and recovery prior to vaccination were 3.8 times more at risk to contract a Covid-19 infection or reinfection in spite of vaccination with at-least one dose of either Covid-19 vaccine.

**Conclusion:** Nearly one in nine HCWs experienced a Covid-19 breakthrough infection in the present study.

## Introduction

Vaccines are considered the mainstay in halting and ending the Covid-19 pandemic which has caused over 172 million cases and 3.7 million deaths worldwide till date [1]. India launched the world’s largest Covid-19 mass vaccination campaign from January’ 2021 in a phased manner beginning with healthcare, sanitation and essential frontline workers, followed by the geriatric population, people with comorbidities, those aged > 45 years and finally the entire adult population [2].

The vaccines approved and deployed in India by the regulatory authority included AZD1222 - ChAdOx1-S (Covishield), manufactured in India by Serum Institute of India through license from Astrazeneca-Oxford [3] and BBV152 (Covaxin), indigenous vaccine developed by Bharat Biotech in collaboration with the Indian Council of Medical Research (ICMR) [4]. The AZD122 (ChAdOx1-S/nCoV-19) recombinant vaccine against Covid-19 is a replication-deficient adenoviral vector vaccine that expresses the SARS-CoV-2 spike protein gene [3]. BBV152 is a whole-virion inactivated SARS-CoV-2 vaccine adjuvanted with Algel-IMDG to induce T-helper-1 cell (Th1) responses [4]. The efficacy of AZD1222 (ChAdOx1-S) after administration of two doses of the vaccines irrespective of interval between the doses has been reported as 63.1%, with possibly higher efficacy on longer intervals [3]. The interim phase-3 clinical trial data reported BBV152 to have efficacy of 78% against infection with SARS-CoV-2 [5]. However, the real-world effectiveness of vaccines may differ from the efficacy reported in clinical trials due to a multitude of factors including the dynamics of disease exposure, diminished antibody response in sub-groups like the elderly and the immunocompromised, and the emergence of newer mutant strains with greater infectivity and virulence [6, 7].

A small proportion of individuals will contract Covid-19 despite complete vaccination as no vaccine accords 100% protection against the disease, and occasionally newer virus variants evolve mechanisms for bypassing the vaccine induced antibody response. Breakthrough infections with reference to Covid-19 refers to the incidence of SARS-CoV-2 infections in individuals who have already been partially or completely vaccinated with any authorized Covid-19 vaccine [8].

According to the ICMR between 0.02 and 0.04% infections have occurred after partial or complete vaccination with either BBV152 or AZD1222 (ChAdOx1-S) [9]. However, healthcare workers (HCWs) represent a very high-risk group for contracting COVID-19 due to sustained occupational exposure to the virus [10, 11], and the breakthrough infection rate assessed in this cohort would provide crucial evidence in understanding the effectiveness of vaccination in preventing symptomatic disease and disease transmission.

The study objective was therefore to determine the breakthrough infection rate of Covid-19 (SARS-CoV-2) infection in those vaccinated with either BBV152 or AZD1222 (ChAdOx1-S) vaccine among healthcare workers.

## Methods

Study design and setting: We conducted a cross-sectional study among healthcare workers of the Maulana Azad Medical College, New Delhi most of whom provide services at the largest dedicated tertiary care Covid-19 hospital in Delhi. As per the government of India policy, two doses of either BBV152 (Covaxin) or AZD1222 (Covishield) vaccine at-least 4 weeks apart were available for administration to all HCWs since January 2021 with no mixing of doses allowed.

The primary outcome of the study was the proportion of breakthrough infection in HCWs which was defined as any Covid-19 infection occurring ≥14 days after receiving both the doses of either of the vaccine(s). The secondary outcome was the proportion of non-breakthrough Covid-19 infections that occurred post vaccination with either a single dose of a Covid-19 vaccine or both doses but prior to a period of 14 days since administration of the second dose.

The independent variables included age, sex, time since vaccination, vaccine type, adherence to non-pharmaceutical measures post-vaccination and previous history of natural Covid-19 infection and recovery prior to vaccination.

The sample size was calculated considering the prevalence of breakthrough infection among HCWs in India as 13.3% as reported in a study from a chronic care facility in Delhi [12], at 95% confidence level, 4% precision, and 10% non-response. The minimum sample size was estimated as 270.

The data was collected during a period of 7 days from May 31 to June 6 through telephonic interviews conducted by multiple trained investigators using the following sources in a consecutive order: (i). Registration records of all the healthcare workers vaccinated at the Covaxin administration site within the college campus during February-March 2021 (ii). Records of the medical interns affiliated to the MAMC who were vaccinated between January-March’ 2021.

### Statistical analysis

Data was entered in Epidata 3.1 (single entered) and analysed with IBM SPSS Version 25 (Armonk, NY). Results were expressed in frequency and proportions for categorical variables, and mean and standard deviation for continuous variables. The significance of difference between proportions was assessed using the chi-square test. A p-value < 0.05 was considered statistically significant.

### Ethics

The study was approved by the Institutional Ethics Committee (F.1/IEC/MAMC/(84/02/2021/No338). Verbal consent was obtained from the HCWs who responded to the telephonic interviews prior to the initiation of the survey.

## Results

We enrolled 325 healthcare workers including 258 medical doctors and interns (79.4%), 52 (16%) frontline health workers, 12 (3.7%) lab technicians and 3 (0.9%) nurses. The mean (SD) age of the participants was 29.1 (9.9) years including 212 (65%) males and 114 (35%) females. Two hundred eighty (90.9%) HCWs were fully vaccinated with two doses while 46 (14.1%) had received only one dose of a Covid-19 vaccine at the time of interview. There were 168 (51.7%) BBV152 recipients and 157 (48.3%) AZD1222 (ChAdOx1-S) recipients. Fifty (15.4%) HCWs reported a past history of natural Covid-19 infection and recovery prior to receiving the first dose of Covid-19 vaccine with 47 (94.1%) being mild cases and 3 (5.9%) being moderate cases needing supplemental oxygen therapy.

A total of 57 (17.5%, 95% C.I 13.8, 22.0) Covid-19 infections comprising 37 breakthrough and 20 non-breakthrough infections were observed in the HCWs vaccinated with at-least one dose of Covid-19 vaccine. The infections were diagnosed with RT-PCR, Ag test, and on clinical suspicion in 48 (84.2%), 3 (5.2%), and 6 (10.5%) cases, respectively. The severity of Covid-19 infections were mild requiring only home isolation in 51 (89.4%) cases while 6 (10.6%) were moderate cases needing supplemental oxygen therapy. Within the households of HCWs suffering Covid-19 infection post-vaccination with at-least one dose of either Covid-19 vaccine, all members were diagnosed concurrently with Covid-19 in 9 (15.8%) cases while at-least one member was infected in 24 (42.1%) cases.

A total of 37 (11.3%, 95% C.I. 8.3, 15.3) breakthrough infections were observed in the HCWs of which 32 (86.5%), 3 (8.1%) and 2 (5.4%) cases were diagnosed with RT-PCR, Ag test and on the basis of clinical symptoms, respectively. The median (IQR) time until incidence of Covid-19 breakthrough infection after receiving the second dose of either Covid-19 vaccine was 47 (28.5, 55) days.

Among the HCWs experiencing a breakthrough infection, 35 (94.6%) were mild cases managed through only home isolation while there were 2 (5.4%) moderate cases requiring supplemental oxygen therapy prior to recovery. Within the households of HCWs reporting the incidence of a Covid-19 breakthrough infection, concurrent infection in all members was observed in 5 (13.5%) cases while at-least one member was infected in 14 (37.8%) cases.

Table 1 summarizes the Covid-19 infection rates in HCWs after administration of at-least a single dose of a Covid-19 vaccine. HCWs without a history of natural Covid-19 infection and recovery prior to vaccination compared to HCWs with such a history were 3.8 times more at risk to contract a Covid-19 infection or reinfection despite vaccination with at-least one dose of either Covid-19 vaccine (p=0.029).

**Table 1.** Distribution of Covid-19 infections post first dose of vaccination in HCWs (N=326)^*^

| Characteristic | Total<br>(N=325) | Total infections*<br>(n=57) | Unadjusted odds<br>95% C.I | p-value |
| --- | --- | --- | --- | --- |
| Vaccination status |  |  |  |  |
| Two doses | 279 (85.8) | 45 (16.1) | 1 | 0.104 |
| One dose | 46 (14.2) | 12 (26.1) | 0.54 (0.26, 1.1) |  |
| Vaccine type |  |  |  |  |
| BBV152 | 168 (51.7) | 33 (19.6) | 0.74 (0.41, 1.3) | 0.303 |
| AZD1222 | 157 (48.3) | 24 (15.3) | 1 |  |
| Age (Years) |  |  |  |  |
| <35 | 255 (78.5) | 46 (18) | 1 | 0.651 |
| ≥35 | 70 (21.5) | 11 (15.7) | 1.2 (0.57, 2.4) |  |
| Sex |  |  |  |  |
| Male | 211 (64.9) | 42 (19.9) | 1 | 0.129 |
| Female | 114 (35.1) | 15 (13.2) | 1.6 (0.86, 3.1) |  |
| HCW Type |  |  |  |  |
| Doctor | 258 (79.4) | 48 (18.6) | 1 | 0.324 |
| Other | 67 (20.6) | 9 (13.4) | 1.4 (0.68, 3.2) |  |
| History of Covid-19 infection<sup>^</sup> |  |  |  |  |
| Present | 50 (15.4) | 3 (6.0) | 3.8 (1.1, 12.7) | 0.029 |
| Absent | 275 (84.6) | 54 (19.6) | 1 |  |
| Masking adherence<sup>+</sup> |  |  |  |  |
| Always | 225 (69.2) | 43 (19.1) | 1 | 0.265 |
| Mostly/other | 100 (30.8) | 14 (14) | 1.4 (0.75, 2.8) |  |
| Social distancing adherence<sup>+</sup> |  |  |  |  |
| Always | 206 (63.4) | 34 (16.5) | 1 | 0.520 |
| Mostly/other | 119 (36.6) | 23 (19.3) | 0.82 (0.46, 1.5) |  |

\*37 breakthrough and 20 non-breakthrough infections
<sup>^</sup>Prior to administration of the first dose of Covid-19 vaccine
<sup>+</sup> Post vaccination with at-least one dose of vaccine

The proportion of breakthrough infections did not show statistically significant variation when compared across subgroups including type of vaccine, age, sex, adherence to non-pharmaceutical measures after vaccination, and history of a natural Covid-19 infection prior to vaccination (Table 2).

**Table 2.** Distribution of factors associated with Covid-19 breakthrough infections in HCWs (N=325)

| Characteristic | Total (N=326) | Breakthrough infection (n=37) | Unadjusted odds 95% C.I | p-value |
| --- | --- | --- | --- | --- |
| Vaccine type |  |  |  |  |
| BBV152 | 168 (51.7) | 24 (14.3) | 1.8 (0.9, 3.8) | 0.092 |
| AZD1222 | 157 (48.3) | 13 (8.3) | 1 |  |
| Age (in Years) |  |  |  |  |
| <35 | 255 (78.5) | 31 (12.2) | 1 | 0.405 |
| ≥35 | 70 (21.5) | 6 (8.6) | 0.67 (0.27, 1.7) |  |
| Sex |  |  |  |  |
| Male | 211 (64.9) | 27 (12.8) | 1 | 0.277 |
| Female | 114 (35.1) | 10 (8.8) | 0.65 (0.3, 1.4) |  |
| HCW Type |  |  |  |  |
| Doctor | 258 (79.4) | 32 (12.5) | 1 | 0.262 |
| Other | 67 (20.6) | 5 (7.5) | 0.57 (0.21, 1.52) |  |
| History of Covid-19 infection* |  |  |  |  |
| Present | 50 (15.4) | 3 (6.0) | 0.45 (0.13, 1.5) | 0.203 |
| Absent | 275 (84.6) | 34 (12.4) | 1 |  |
| Masking adherence<sup>+</sup> |  |  |  |  |
| Always | 225 (69.2) | 26 (11.6) | 1 | 0.884 |
| Mostly/other | 100 (30.8) | 11 (11.0) | 0.94 (0.45, 1.9) |  |
| Social distancing adherence<sup>+</sup> |  |  |  |  |
| Always | 206 (63.4) | 21 (10.2) | 1 | 0.375 |
| Mostly/other | 119 (36.6) | 16 (13.4) | 1.4 (0.68, 2.7) |  |

\*Prior to vaccination; + Post vaccination with at-least one dose of vaccine

## Discussion

In this study, nearly, one in five HCWs reported the incidence of Covid-19 infection after receiving at-least one dose of a Covid-19 vaccine but prior to either complete vaccination or before 14 days post administration of the second vaccine dose. Furthermore, one in nine HCWs experienced a breakthrough infection after being administered both scheduled doses of either Covid-19 vaccine. These findings suggest that in real world settings a significant proportion of vaccinated individuals with high risk of exposure remain vulnerable to Covid-19 infection albeit with reduced disease severity in most cases.

Vaccine seroconversion through robust anti-spike antibody response is likely to be induced after a single dose of AZD1222 (ChAdOx1-S) compared to BBV152 wherein two doses are usually required to stimulate adequate antibody levels [7]. However, in the present study, although breakthrough and non-breakthrough infections were higher in the BBV152 group compared to the AZD1222 group, the differences were not statistically significant.

Evidence from a previous study suggests that a single dose of either BBV152 or AZD1222 induced higher concentration of neutralizing IgG antibody in those having a history of natural infection and recovery from COVID-19 [13]. Similarly, in this study, a history of a natural infection and recovery from Covid-19 was observed to be protective against a subsequent Covid-19 infection or reinfection in those HCWs who had been administered at-least a single dose of Covid-19 vaccine.

The rates of breakthrough infection observed in the present study (11.3%) were slightly lower than that observed during surveillance in a chronic care facility in Delhi, India (13.2%) where HCWs received either AZD1222 (ChAdOx1-S) or BBV152 [12]. However, another study in a large cohort of HCWs from a north Indian city vaccinated with AZD1222 (ChAdOx1-S) reported the incidence of Covid-19 breakthrough infections to be only 1.6% (48 out of 3,000) while 2.6% tested positive after receiving at-least one dose of the vaccine [14]. In contrast, Hacisuleyman et al. report the incidence of breakthrough infection as just 0.5% in a cohort of 417 health care workers who had previously received two doses the BNT162b2 (Pfizer– BioNTech) or mRNA-1273 (Moderna) vaccine [15].

The period of observation in this study coincided with a massive wave of the Covid-19 epidemic in Delhi during April and May 2021 which witnessed 0.737 million cases including 11,075 deaths [16]. Furthermore, the emergent evidence from genomic analysis also reveals that the Covid-19 variants of concern, B.1.617.2 (Delta) and B.1.1.7 (Alpha) having ∼50% higher transmissibility were primarily responsible for the surge in cases during the same period. These variants also constituted the predominant lineages found in the breakthrough infections cases due to a probable immune escape mechanism that could occasionally bypass the vaccine induced immunity [17]. Moreover, diminished neutralizing antibody activity and limited protectiveness against the delta variant of the SARS-CoV-2 has been observed in most of the currently available Covid-19 vaccines globally especially prior to complete vaccination with both vaccine doses [18, 19].

The strengths of the study are that it was conducted in real-world settings with the period of observation inclusive of the peak of the Covid-19 pandemic in Delhi, India when health systems were overwhelmed resulting in large scale viral exposure of HCWs providing either outpatient or inpatient treatment services. However, there are certain study limitations. First, since infection status of the HCWs was based on self-report in the absence of a mandatory testing policy, mostly symptomatic breakthrough infections diagnosed with RT-PCR or the Ag test were likely to be captured while asymptomatic infections also capable of viral transmission were potentially omitted. Second, although, we observed high rates of transmission in the household members of the HCWs, the vaccination status in the infected household members was not recorded. Third, non-breakthrough infection rate could be inflated since the HCWs at the time of administration of their first dose of vaccine may already have been infected. Fourth, comorbidity status was not ascertained in the participants although the likelihood of underlying morbidities in the HCWs was less considering their low median age. Consequently, future studies should assess the extent of vaccine induced antibody response and protection after vaccination with Covid-19 vaccines in patients with diabetes, heart disease, chronic kidney disease, and older people at risk of reduced protection in addition to pre-existing concerns causally linked with the occurrence of severe disease. A final limitation is that the sample size was not adequately powered to detect statistically significant differences between subgroups.

In conclusion, breakthrough infections represent a major public health challenge in ending the Covid-19 pandemic. Robust surveillance through large-scale epidemiological studies to identify the predictors of breakthrough infection among individuals at risk, and rapid genomic analysis for early recognition of emerging variants of concern that have greater capability of causing breakthrough infections warrant continued prioritization.

## Data Availability

The anonymized dataset will be made available on reasonable request from the corresponding author.

## Sources of support

Nil

## Conflicts of interest

None

## Notes

### Competing Interest Statement

The authors have declared no competing interest.

### Author Declarations

Institutional Ethics Committee, Maulana Azad Medical College & Associated Hospitals. (F.1/IEC/MAMC/(84/02/2021/No338)

### Summary of Updates

1. Non-breakthrough infections were defined in the methods section 2. Non-breakthrough infections with symptomatic history were verified and cross-checked, and only those with strong clinical suspicion of Covid-19 were retained as per the operational definition. Consequently, the total number of non-breakthrough infections reduced to

